# Differential response to deep brain stimulation in harm-avoidant vs. disgust subtypes of obsessive-compulsive disorder

**DOI:** 10.64898/2026.07.20.26357997

**Authors:** Thomas Hamre, Thomas Kutcher, Rick Hanish, Sarah Soubra, Holden Bentley, Kasra Mansourian, Grace Nitcheu, Vinayak Belavadi, Saipravallika Chamarthi, Zahra Jourahmad, Mohammed Hasen, Nisha Giridharan, Sarah R. Heilbronner, Eric A. Storch, Wayne K. Goodman, Nicole R. Provenza, Sameer A. Sheth

## Abstract

**Background:** Deep brain stimulation (DBS) is a promising intervention for obsessive-compulsive disorder (OCD) refractory to conventional therapies. However, preoperative factors that reliably predict treatment response are not well established. Among patients with contamination OCD, some are concerned with substances potentially causing harm to themselves or others (“harm-avoidant”), while others are viscerally repulsed by them (“disgust”). Disgust-oriented OCD is associated with attenuated psychotherapy outcomes; relationship with DBS outcome has yet to be examined. Here, we compare clinical outcomes following DBS between harm-avoidant and disgust OCD subtypes.

**Methods:** We conducted a retrospective cohort study of 14 contamination OCD patients (7 female, 7 male) undergoing DBS between 2019 and 2024 and categorized them as harm-avoidant or disgust based on clinical features. We assessed demographics and Yale-Brown Obsessive-Compulsive Scale-Second Edition (Y-BOCS-II) scores preoperatively and longitudinally.

**Results:** Among 14 patients with contamination OCD, the harm-avoidant (n=10) and disgust (n=4) groups had similar demographic characteristics and baseline Y-BOCS-II scores (harm-avoidant: 40.4 ± 4.2, disgust: 40.0 ± 6.9, *p*=0.92, Welch’s t-test). At the latest follow-up, harm-avoidant patients achieved significantly lower Y-BOCS-II scores (harm-avoidant: 13.8 ± 11.1, disgust: 30.5 ± 3.1; *p*=0.001, Welch’s t-test). Nine of ten harm-avoidant patients achieved response status (≥35% score reduction) compared to only one of four disgust patients (*p*=0.04, Fisher’s exact test).

**Conclusions:** Disgust OCD patients experienced attenuated improvement following DBS compared to harm-avoidant OCD patients. If confirmed in larger cohorts, these findings could motivate preoperative phenotypic stratification, inform decision-making about DBS candidacy, and guide alternative therapeutic strategies for disgust patients.

## INTRODUCTION

Obsessive-compulsive disorder (OCD) is a potentially debilitating neuropsychiatric condition marked by intrusive thoughts and repetitive behaviors, affecting 1-2% of the global population [1]. While first-line treatments such as exposure and response prevention (ERP) therapy and selective serotonin reuptake inhibitors (SSRIs) are effective for many OCD patients, 10-20% do not respond to these initial interventions [2,3]. For this treatment-resistant population, deep brain stimulation (DBS) targeting the ventral capsule/ventral striatum (VC/VS) has proven effective, with some studies reporting response rates exceeding 60% [4–6].

Although DBS is effective for many OCD patients, preoperative predictors of treatment response remain elusive [7–8]. Limited meta-analytic evidence suggests that later OCD onset and sexual or religious obsessions may be associated with better DBS response, whereas hoarding symptoms had been linked to poorer DBS response before its reclassification in DSM-5 as a separate disorder. These few phenotypic predictors are a start, but greater understanding of subtypes that are more or less likely to respond to this therapy would be very useful for clinical decision-making and prognostication [2,4,7–9]. In contrast, the well-established relationship between Parkinson’s disease (PD) phenotypes and the therapeutic effects of DBS has led to significantly improved response rates over time for PD patients receiving DBS [10–12]. For example, clinicians can reliably predict favorable outcomes after DBS in patients with a tremor-dominant PD phenotype [13–14]. Conversely, patients with PD characterized by postural instability and gait disturbances tend to respond less favorably to DBS [15]. The ability to similarly identify which OCD patients will benefit most from DBS therefore represents a critical unmet need. OCD comprises at least four distinct subtypes: (1.) contamination obsessions with cleaning compulsions, (2.) symmetry obsessions with ordering or counting, (3.) doubt and harm obsessions with checking behaviors, and (4.) intrusive taboo thoughts with neutralizing rituals, yet predictors of response based on these subtypes have not emerged [16].

While these traditional OCD subtypes do not appear to influence DBS response, other distinctions in OCD presentation may. Emerging research supports dividing contamination-OCD patients into two groups: those with symptoms driven by harm-avoidance versus disgust [17,18]. Patients with a harm-avoidant phenotype avoid perceived contaminants to prevent feared future outcomes, such as contracting an illness. In contrast, patients with a disgust phenotype experience intense, immediate feelings of revulsion when exposed to contaminants. These evoked disgust sensations trigger cleaning rituals independent of perceived risk of harm. Since disgust responses in both healthy individuals and those with psychiatric disorders tend to be resistant to ERP therapy, a key treatment for OCD, this distinction already holds clinical relevance [19–22].

Phenotypic classification into harm-avoidant or disgust contamination OCD phenotypes may plausibly influence DBS outcomes. In OCD, a malfunctioning threat detection system is thought to overestimate the risk of negative outcomes and promote defensive responses, a dysfunction mediated by hyperactivity within cortico-striato-thalamo-cortical (CSTC) circuits [9, 23–26]. VC/VS DBS directly modulates these circuits, and the harm-avoidant contamination phenotype, in which compulsions serve to avert a feared consequence, aligns directly with this traditional model of OCD pathophysiology. In contrast, patients with a disgust phenotype engage in neutralizing behaviors not to avert a feared outcome but to eliminate the feeling of disgust itself, a state closely linked to activation of the anterior insula, a region remote from the CSTC circuitry modulated by VC/VS DBS [27, 28]. Clinically, disgust is recognized as a contributor to exposure and response prevention (ERP) non-response and is generally more resistant to extinction than fear [29]. DBS and ERP appear to act synergistically, implying a shared mechanism, so the features that blunt ERP response in disgust-contamination OCD may also blunt VC/VS DBS response [30, 31]. We therefore retrospectively analyzed DBS outcomes in a cohort of contamination OCD patients, comparing those with harm-avoidant and disgust phenotypes.

## METHODS

We conducted a retrospective cohort study to evaluate clinical outcomes in OCD patients who underwent DBS at our institution between 2019 and 2024. All included patients had a documented minimum of a five-year history of OCD, failed at least three SSRI trials (including at least one with antipsychotic augmentation), completed at least 20 ERP therapy sessions, and had a baseline Y-BOCS-II > 28 before undergoing DBS. From this full cohort of OCD patients who received DBS, we identified those with the contamination OCD subtype and further classified them into harm-avoidant or disgust groups through a comprehensive review of psychiatric electronic medical records describing each patient’s OCD symptomatology. Our expert psychiatrist (WG) and psychologist (ES) verified all diagnostic classifications for accuracy.

We collected demographic information, including disease duration, age at DBS implantation, and psychiatric comorbidities. We assessed clinical outcomes using the Yale-Brown Obsessive-Compulsive Scale-Second Edition (Y-BOCS-II) and the Hamilton Depression Rating Scale (HAM-D), with scores recorded before surgery and at the most recent follow-up after DBS [32,33]. For longitudinal analysis, we gathered Y-BOCS-II and HAM-D scores from each follow-up visit. Two patients in the harm-avoidant group were never administered the HAM-D scale and were therefore excluded from HAM-D analyses. Statistical analyses included Welch’s t-test for continuous variables, paired t-tests for within-group comparisons, and Fisher’s exact test for binary outcomes. We conducted all analyses with Python 3.13.

At our center, the neurosurgical team uses a frame-based, robot-assisted workflow for VC/VS DBS implantation. High-resolution preoperative magnetic resonance imaging and computed tomography scans are merged on a ROSA planning station (Zimmer Biomet). The anterior commissure to posterior commissure line and the midsagittal interhemispheric plane are then marked, and two bilateral trajectories are planned: an anterior VC/VS path at the junction of the anterior commissure and the anterior limb of the internal capsule, and a posterior path toward the bed nucleus of the stria terminalis lateral to the intersection of the anterior commissure and the fornix. The deepest contact on each trajectory is placed at the white matter-to-gray matter junction adjacent to the ventral striatum, using contrast imaging to avoid blood vessels. Intraoperatively, after Leksell G frame placement and O-arm (Medtronic) computed tomography registration via frame pins, a single burr hole on each side accommodates both trajectories. Intraoperative awake testing involves a rapid monopolar review of all four contacts on each lead; if positive responses (such as improved mood or reduced anxiety) occur, the lead is secured. Otherwise, the cannula is redirected to the alternate target. Stimulation usually begins around 2 weeks after surgery [34].

## RESULTS

From an initial cohort of 22 OCD patients who underwent VC/VS DBS, we included 14 with contamination OCD, ten of whom exhibited primarily harm-avoidant traits and four disgust traits. One disgust patient had 581 days of postoperative follow-up; all other included patients had more than two years of follow-up. Mean follow-up duration did not differ significantly between groups (harm-avoidant: 1149.0 ± 338.4 days, disgust: 1168.8 ± 692.8 days, *p*=0.96, Welch’s t-test). Demographic data and clinical outcome measures for both groups are summarized in Table 1. The harm- and disgust groups showed no significant differences in mean disease duration (harm-avoidant: 18.3 ± 9.2 years, disgust: 19.8 ± 3.1 years, *p*=0.66, Welch’s t-test), age at DBS intervention (harm-avoidant: 36.7 ± 15.0 years, disgust: 32.8 ± 8.3 years, *p*=0.55, Welch’s t-test), or baseline Y-BOCS-II scores (harm-avoidant: 40.4 ± 4.2, disgust: 40.0 ± 6.9, *p*=0.92, Welch’s t-test).

**Table 1.** Demographic and clinical data for harm-avoidant and disgust contamination OCD patients.

|  | Harm-Avoidant | Disgust | *p*-value |
| --- | --- | --- | --- |
| No. of patients | 10 | 4 |  |
| F/M sex, n | 6/4 | 1/3 |  |
| Comorbidities, n (%) |  |  |  |
| GAD | 5 (50%) | 2 (50%) |  |
| MDD | 6 (60%) | 2 (50%) |  |
| Tourette Syndrome | 1 (10%) | 1 (25%) |  |
| Disease duration, yrs | 18.3 ± 9.2 | 19.8 ± 3.1 | 0.66 |
| Age at surgery, yrs | 36.7 ± 15.0 | 32.8 ± 8.3 | 0.55 |
| Baseline Y-BOCS-II score | 40.4 ± 4.2 | 40.0 ± 6.9 | **0.92** |
| Follow-up Y-BOCS-II score | 13.8 ± 11.1 | 30.5 ± 3.1 | **0.001** |
| Y-BOCS score improvement, % | 66.3% ± 25.2% | 21.7% ± 17.2% | **0.005** |
| Follow-up duration, d | 1149.0 ± 338.4 | 1168.8 ± 692.8 |  |
| ≥35% Y-BOCS-II reduction, n (%) | 9 (90%) | 1 (25%) | **0.04** |
| Baseline HAMD score | $17.8 \pm 7.8$ | $18.5 \pm 6.0$ | 0.86 |
| Follow-up HAMD score | $6.6 \pm 8.2$ | $19.0 \pm 4.1$ | **0.006** |
| HAMD score improvement,<br>% | $57.3\% \pm 40.9\%$ | $-15.9\% \pm 63.3\%$ | 0.098 |
| Follow-up duration (HAM-D<br>subgroup), d | $1040.1 \pm 562.6$ | $1168.8 \pm 692.8$ | |

At the most recent follow-up, the harm-avoidant group had significantly lower Y-BOCS-II scores (13.8 ± 11.1) than the disgust group (30.5 ± 3.1, *p*=0.001, Welch’s t-test, Figure 1A, B). Likewise, the percentage reduction in Y-BOCS-II scores from each patient’s preoperative baseline to the latest follow-up was significantly greater in the harm-avoidant group (66.3% ± 25.2%) than in the disgust group (21.7% ± 17.2%, *p*=0.005, Welch’s t-test, Figure 1C, D). We observed a similar pattern with HAM-D scores (harm-avoidant: n=8, disgust: n=4). Despite comparable baseline means between groups (harm-avoidant: 17.8 ± 7.8, disgust: 18.5 ± 6.0, p=0.86, Welch’s t-test), HAM-D scores for the disgust group were significantly higher than those of the harm-avoidant group at the most recent follow-up (harm-avoidant: 6.6 ± 8.2, disgust: 19.0 ± 4.1, p=0.006, Welch’s t-test, Figure 2).

**Figure 1.**
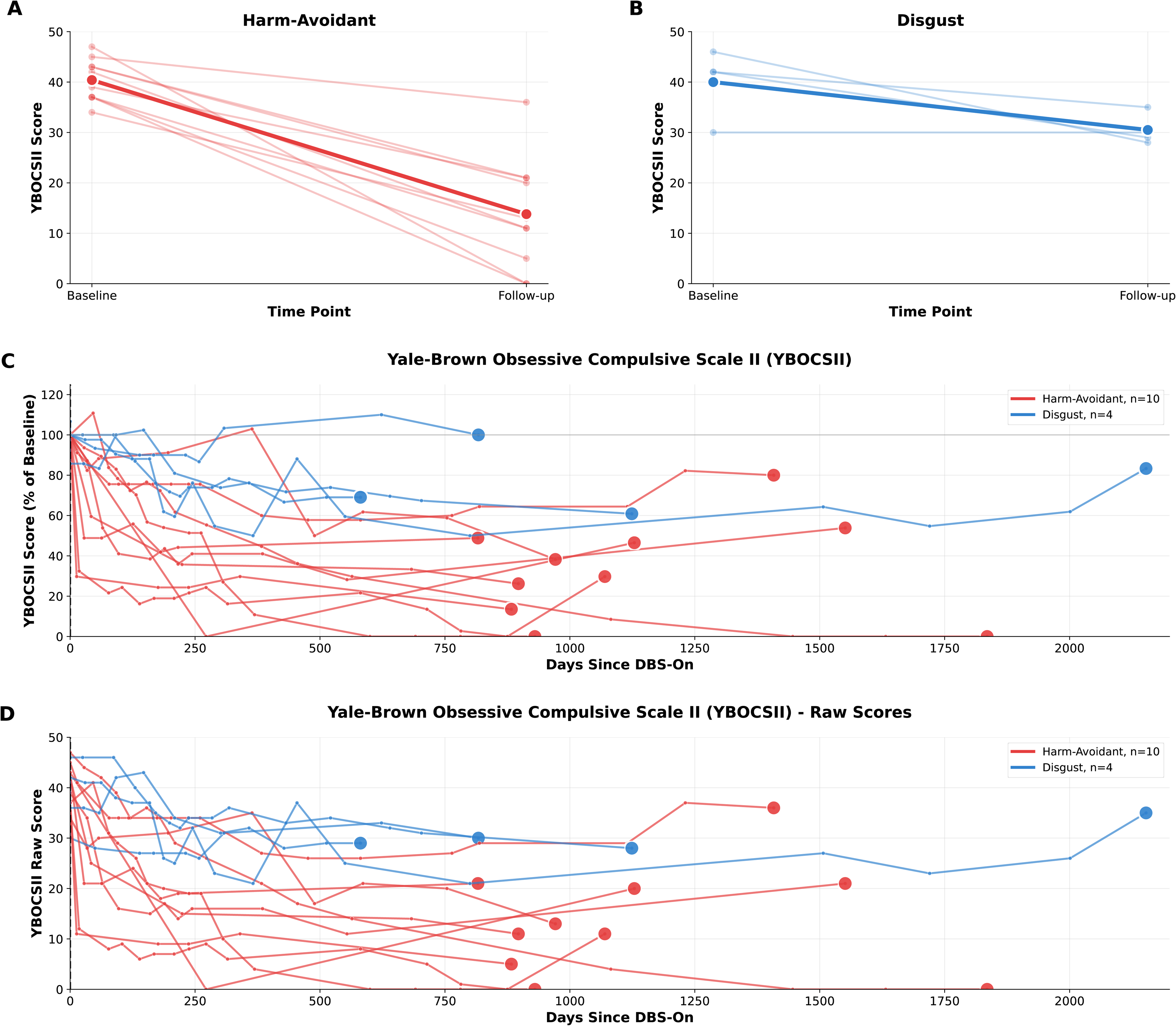
Clinical response to ventral capsule/ventral striatum DBS in harm-avoidant (n=10) versus disgust (n=4) contamination OCD patients. (A–B) Y-BOCS-II scores before and after DBS: harm-avoidant showed a significant within-group reduction (*p*<0.001, paired t-test), whereas disgust did not (*p*=0.09, paired t-test). (C–D) Longitudinal trajectories (normalized to each patient’s baseline in C; raw scores in D) illustrate persistently higher symptom severity in the disgust cohort; the between-group difference at the latest follow-up favored the harm-avoidant group (*p*=0.001, Welch’s t-test).

**Figure 2.**
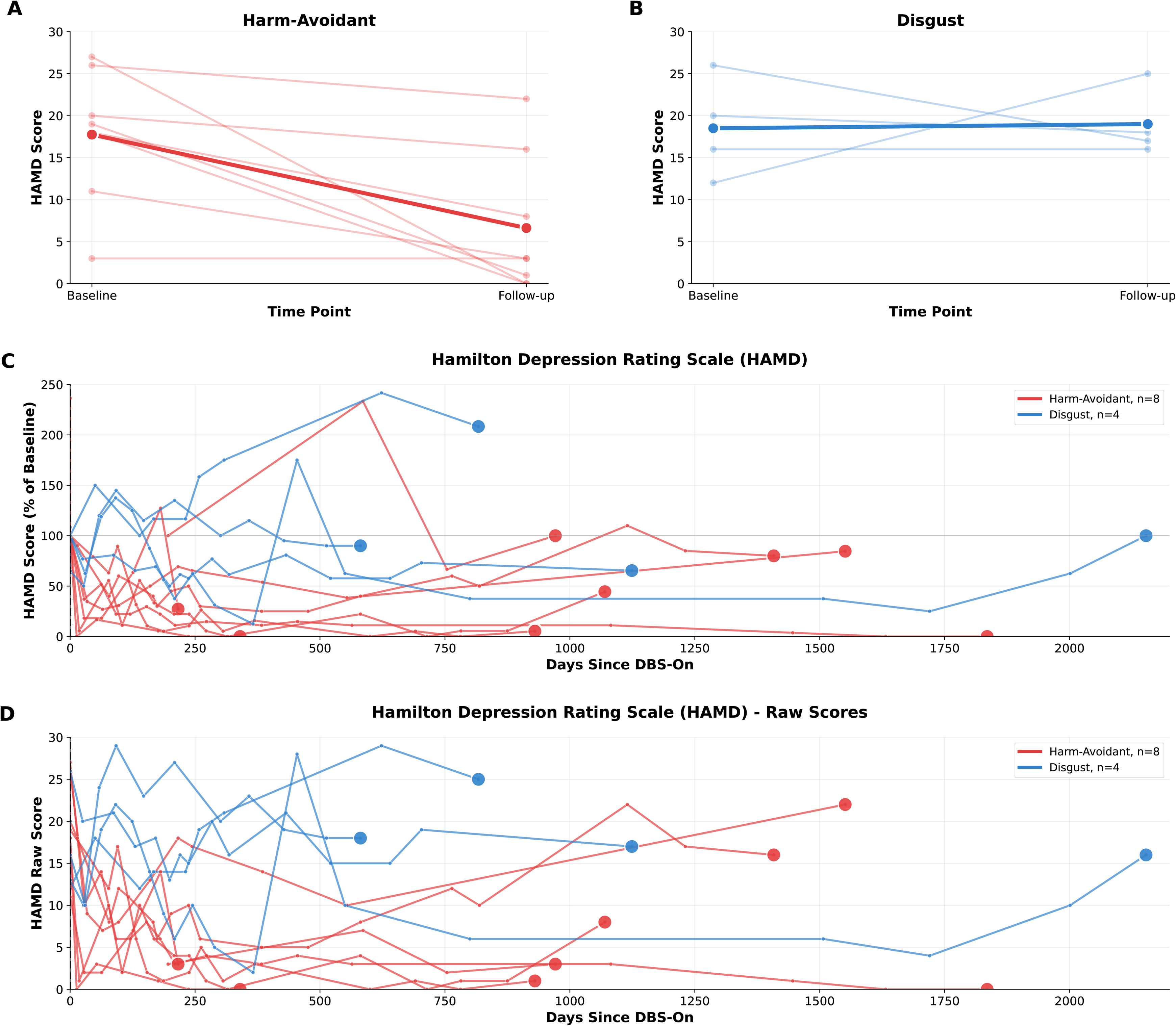
Hamilton Depression Rating Scale (HAM-D) scores over time following DBS in harm-avoidant (n=8) versus disgust (n=4) contamination OCD patients. (A–B) Pre- versus post-DBS HAM-D scores are displayed for each group. (C–D) Longitudinal trajectories (normalized in C; raw scores in D) reveal a higher depressive symptom burden in the disgust cohort. The between-group difference at the latest follow-up was significant (*p*=0.006, Welch’s t-test), whereas baseline group means did not differ (*p*=0.86, Welch’s t-test). Note: OCD = obsessive-compulsive disorder; DBS = deep brain stimulation; GAD = generalized anxiety disorder; MDD = major depressive disorder; Y-BOCS-II = Yale-Brown Obsessive Compulsive Scale, Second Edition; HAM-D = Hamilton Depression Rating Scale. HAM-D scales were not administered to two harm-avoidant patients; for those analyses, eight harm-avoidant patients are compared with four disgust patients.

Within groups, only the harm-avoidant group showed a significant reduction in Y-BOCS-II scores from baseline (40.4 ± 4.2) to the most recent follow-up (13.8 ± 11.1, *p*<0.001, paired t-test). For the disgust group, the change in Y-BOCS-II scores from baseline (40.0 ± 6.9) to the latest follow-up (30.5 ± 3.1, *p*=0.09, paired t-test) did not achieve significance. Defining “responder status” as a ≥35% reduction in Y-BOCS-II scores from pre-DBS baseline, nine of ten harm-avoidant patients achieved responder status, compared with only one of the four disgust patients (*p*=0.04, Fisher’s exact test).

## DISCUSSION

In this retrospective cohort study of 14 patients with severe, treatment-resistant contamination OCD, those with disgust contamination OCD experienced significantly less improvement after DBS compared to those with harm-avoidant presentations. Prior meta-analytic work has documented symptom-level heterogeneity in DBS response, identifying patients with hoarding phenotypes as comparatively poor DBS responders. This finding, together with the subsequent reclassification of hoarding as a distinct disorder in DSM-5, implicates neurobiologically dissociable OCD subtypes with differential sensitivity to neuromodulation [4]. The differential response to DBS based on contamination OCD phenotype can be understood through an approach-avoidance framework of OCD. The so-called ‘approach system,’ mediated by dopaminergic pathways projecting from the ventral tegmental area to the nucleus accumbens (ventral striatum) and to frontal cortical regions, including the ventromedial prefrontal cortex, promotes goal-oriented behaviors and reinforces actions leading to rewarding outcomes [35,36]. Conversely, an ‘avoidance system’ composed of circuits centered on amygdala-periaqueductal gray pathways, with the dorsal anterior cingulate cortex and other frontal regions involved in assessing conflict and anticipating aversive outcomes, coordinates avoidant or defensive behaviors [37,38]. When functioning correctly, these complementary systems facilitate flexible, appropriate shifts between approach and avoidance behaviors in response to environmental cues. In OCD, this balance is fundamentally disrupted, with CSTC circuits exhibiting characteristic hyperactivity in the lateral orbitofrontal cortex and anterior cingulate, along with altered connectivity patterns that promote behavioral inflexibility [39–42]. Consequently, excessive and costly avoidance behaviors are driven by an imbalance between goal-directed systems and habitual control and avoidance systems [43,44].

VC/VS DBS can generally be understood as treating OCD by modulating CSTC circuits to restore the brain’s ability to perform flexible, goal-directed behavior. It achieves this effect in part by disrupting a pathologically rigid network state that drives compulsive avoidance [25, 45–47]. We have posited that DBS of this region increases reward sensitivity, thereby facilitating pro-approach behavior. Behavioral evidence for this hypothesis includes the induction of approach behaviors such as smiling, talkativeness, extroversion, and energy immediately upon stimulating this region, even acutely in the operating room [34]. These behaviors are evoked whether the primary diagnosis is OCD [25, 34, 47–49] or depression [50,51]. As connectomic evidence, studies using diffusion tractography show that DBS of this region engages a fronto-basal ganglia control pathway, including fibers originating from medial frontal areas, such as the dorsal anterior cingulate cortex and the pre-supplementary motor area, which are involved in conflict monitoring and cognitive control, along with orbitofrontal cortex regions linked to outcome valuation and reward-oriented behavior [52–54]. These fibers pass through the anterior limb of the internal capsule to reach the ventral striatum and the limbic/anterior subthalamic nucleus, forming a circuit well-positioned to regulate fear/threat-driven avoidance behaviors and promote goal/reward-motivated behaviors.

During symptom provocation, VC/VS stimulation reduces pathologically elevated coupling between the nucleus accumbens and medial prefrontal and orbitofrontal threat-assessment regions, allowing for more flexible engagement of goal-directed striatal circuits [55]. Electrophysiological data from chronic intracranial neural recordings also show that DBS modulates low-frequency VS neural variability, increasing it from a pathologically low degree of variability in the symptomatic state to a healthy amount in the state of clinical response [25]. Taken together, VC/VS stimulation appears to modulate threat-assessment and cognitive-control networks that drive compulsive avoidance and facilitate reward-sensitive pro-approach behavior. In individuals with OCD, this approach-oriented behavior may promote learning of new, non-threatening relationships with previously provoking stimuli and thus gradually lead to greater capability to resist pathological avoidant behavior.

Emerging psychiatric research supports the distinction between harm-avoidant and disgust contamination OCD symptoms, a distinction linked to partially independent circuits responsible for these two phenotypes [17,18,28,56]. The harm-avoidant subtype would be expected to respond more readily to therapies that preferentially modulate the approach-avoidance axis, such as ERP and VC/VS DBS [1,57–59]. This expectation aligns with the results observed in our cohort, in which patients exhibiting the harm-avoidant phenotype responded much more robustly. In contrast, disgust contamination OCD symptoms appear to arise primarily from an inappropriate “neural reuse” of interoceptive and viscerosensory networks centered on the anterior insula that process revulsion, nausea, and bodily salience [60,61]. Because these symptom manifestations do not originate from avoidance due to misjudgment of potential threat, stimulation that promotes pro-approach behaviors by modulating threat-assessment and cognitive-control circuits may be less effective.

Our observation that disgust contamination OCD shows a reduced response to VC/VS DBS has several implications. First, if confirmed in larger samples, this subtype may be one to avoid or at least to counsel appropriately when determining DBS candidacy. Developing and validating scales that more accurately measure the disgust component contributing to OCD symptoms could be necessary to support such subtyping. Second, patients with disgust symptoms receiving DBS for OCD might benefit from targeted adjunctive treatments, such as cognitive reappraisal or interoceptive-exposure therapy, that more directly address the unique basis of their contamination symptoms [62,63].

Several notable limitations affect the interpretation of our results. The small sample size in both patient groups may limit the generalizability of our findings to larger populations. In particular, the disgust group was markedly smaller than the harm-avoidant group; without detailed incidence data from larger cohorts, it remains unclear whether this imbalance reflects biased sampling. Additionally, phenotypic classification was subjective, based on a retrospective review of electronic medical records with expert verification, rather than the consistent use of objective disgust-assessment scales. This harm-avoidant and disgust distinction may also oversimplify a more complex phenotypic difference. Medication adjustments, varying levels of participation in exposure therapy, comorbid affective symptoms, and ongoing adjustments of stimulation parameters during follow-up visits after implantation could also confound outcomes. Nonetheless, the pattern we observe, substantial OCD symptom reduction in harm-avoidant contamination OCD presentations and relatively modest changes in disgust presentations, aligns with a broader body of behavioral, neuroimaging, and neurophysiological evidence suggesting that avoidant phenotypes in OCD may be the most responsive to VC/VS stimulation.

## CONCLUSION

Patients with disgust contamination OCD exhibit significantly less improvement with VC/VS DBS compared to those with harm-avoidant contamination OCD, despite having similar baseline disease severity. These results suggest that disgust processing may be less directly modulated by VC/VS stimulation and align with the developing hypothesis that VC/VS DBS facilitates pro-approach behavior and thus more successfully treats avoidant DBS subtypes. Identifying OCD subtypes with lesser or greater responsiveness to DBS would aid candidacy evaluation, guide patient counseling, and potentially inform alternative neuromodulation targets or treatment modalities for patients with unfavorable disease phenotypes.

## Data Availability

All data produced in the present study are available upon reasonable request to the authors.

## Funding

This work was supported by the National Institutes of Health (NIH) National Institute of Neurological Disorders and Stroke (NINDS) BRAIN Initiative [grant number UH3NS100549] and The McNair Foundation. The study sponsors had no role in study design, data collection, analysis, interpretation, or manuscript preparation.

## Author contributions (CRediT)

T.H., N.R.P., W.K.G., and S.A.S. conceptualized and designed the study; T.H., T.K., and R.H. curated the clinical data; E.A.S. and W.K.G. collected the clinical data and verified the diagnostic (harm-avoidant vs. disgust) classifications; T.H. performed the statistical analysis; T.H. drafted the manuscript; S.S., H.B., K.M., G.N., V.B., S.C., Z.J., M.H., N.G., and S.R.H. critically revised the manuscript for important intellectual content; and all authors approved the final version.

## Data and code availability

The analysis code and de-identified data supporting these findings are available from the corresponding author upon reasonable request, subject to institutional data-sharing agreements.

This work has not been previously published or posted as a preprint.

## DECLARATION OF COMPETING INTEREST

Dr. Sheth is a consultant for Boston Scientific, Zimmer Biomet, Abbott, Koh Young, and Neuropace. He is co-founder of Motif Neurotech. Dr. Sheth and Dr. Provenza report receiving funding grants from the National Institutes of Health (NIH) National Institute of Neurological Disorders and Stroke BRAIN Initiative and The McNair Foundation. Dr. Storch reports receiving research funding to his institution from the Ream Foundation, International OCD Foundation, Wellcome Trust, MHNTI, and NIH. He receives direct funding from the International OCD Foundation as well as MHNTI for providing trainings on treating obsessive-compulsive disorder with psychotherapy. He was a consultant for Brainsway and Biohaven Pharmaceuticals in the past 36 months. He owns stock options less than $5000 in NView (for distribution of the Y-BOCS and CY-BOCS) and receives royalties from OCD Scales LLC (for distribution of the Y-BOCS and CY-BOCS). He receives book royalties from Elsevier, Wiley, Oxford, American Psychological Association, Guildford, Springer, Routledge, and Jessica Kingsley. Dr. Goodman receives royalties from Nview, LLC and OCDscales, LLC. The other authors have no personal, financial, or institutional interest in any of the drugs, materials, or devices described in this article.

## Notes

### Author Declarations

This retrospective study was approved by the Baylor College of Medicine Institutional Review Board (protocols H-53761 BUILDING MOOD STATE CLASSIFIERS TO INFORM DEEP BRAIN STIMULATION OF TREATMENT-RESISTANT BIPOLAR DEPRESSION and H-56119 MONITORING IN PATIENTS THAT HAVE UNDERGONE DEEP BRAIN STIMULATION (DBS) FOR NEUROPSYCHIATRIC DISORDERS) and was conducted in accordance with the Declaration of Helsinki. All patients had provided written informed consent for deep brain stimulation and for the research use of their clinical data; capacity to provide informed consent was assessed by the treating clinical team.

## REFERENCES

1. Goodman WK, Storch EA, Sheth SA (2021): Harmonizing the Neurobiology and Treatment of Obsessive-Compulsive Disorder. Am J Psychiatry 178:17–29.

2. Hirschtritt ME, Bloch MH, Mathews CA (2017): Obsessive-Compulsive Disorder: Advances in Diagnosis and Treatment. JAMA 317:1358–1367.

3. Raviv N, Staudt MD, Rock AK, MacDonell J, Slyer J, Pilitsis JG (2020): A Systematic Review of Deep Brain Stimulation Targets for Obsessive Compulsive Disorder. Neurosurgery 87:1098.

4. Gadot R, Najera R, Hirani S, Anand A, Storch E, Goodman WK, et al. (2022): Efficacy of deep brain stimulation for treatment-resistant obsessive-compulsive disorder: systematic review and meta-analysis. J Neurol Neurosurg Psychiatry jnnp-2021–328738.

5. Graat I, Mocking R, Figee M, Vulink N, de Koning P, Ooms P, et al. (2021): Long-term Outcome of Deep Brain Stimulation of the Ventral Part of the Anterior Limb of the Internal Capsule in a Cohort of 50 Patients With Treatment-Refractory Obsessive-Compulsive Disorder. Biol Psychiatry 90:714–720.

6. Sheth SA, Provenza NR, Soubra S, Hamre T, Shofty B, Banks GP, et al. (2026): Early feasibility study of sensing-enabled ventral capsule deep brain stimulation in 10 participants with intractable obsessive-compulsive disorder. medRxiv.

7. Alonso P, Cuadras D, Gabriëls L, Denys D, Goodman W, Greenberg BD, et al. (2015): Deep Brain Stimulation for Obsessive-Compulsive Disorder: A Meta-Analysis of Treatment Outcome and Predictors of Response. PLoS One 10:e0133591.

8. Martinho FP, Duarte GS, Couto FSD (2020): Efficacy, Effect on Mood Symptoms, and Safety of Deep Brain Stimulation in Refractory Obsessive-Compulsive Disorder: A Systematic Review and Meta-Analysis. J Clin Psychiatry 81:19r12821.

9. Dougherty DD, Brennan BP, Stewart SE, Wilhelm S, Widge AS, Rauch SL (2018): Neuroscientifically Informed Formulation and Treatment Planning for Patients With Obsessive-Compulsive Disorder: A Review. JAMA Psychiatry 75:1081–1087.

10. Armstrong MJ, Okun MS (2020): Diagnosis and Treatment of Parkinson Disease: A Review. JAMA 323:548–560.

11. Hariz M, Blomstedt P (2022): Deep brain stimulation for Parkinson’s disease. J Intern Med 292:764–778.

12. Ravi DK, Baumann CR, Bernasconi E, Gwerder M, Ignasiak NK, Uhl M, et al. (2021): Does Subthalamic Deep Brain Stimulation Impact Asymmetry and Dyscoordination of Gait in Parkinson’s Disease? Neurorehabil Neural Repair 35:1020–1029.

13. Hitti FL, Ramayya AG, McShane BJ, Yang AI, Vaughan KA, Baltuch GH (2019): Long-term outcomes following deep brain stimulation for Parkinson’s disease. J Neurosurg 132:205–210.

14. Kremer NI, Pauwels RWJ, Pozzi NG, Lange F, Roothans J, Volkmann J, Reich MM (2021): Deep Brain Stimulation for Tremor: Update on Long-Term Outcomes, Target Considerations and Future Directions. J Clin Med 10:3468.

15. Abboud H, Genc G, Thompson NR, Oravivattanakul S, Alsallom F, Reyes D, et al. (2017): Predictors of Functional and Quality of Life Outcomes following Deep Brain Stimulation Surgery in Parkinson’s Disease Patients: Disease, Patient, and Surgical Factors. Parkinsons Dis 2017:5609163.

16. Bloch MH, Landeros-Weisenberger A, Rosario MC, Pittenger C, Leckman JF (2008): Meta-Analysis of the Symptom Structure of Obsessive-Compulsive Disorder. Am J Psychiatry 165:1532–1542.

17. Melli G, Bulli F, Carraresi C, Tarantino F, Gelli S, Poli A (2017): The differential relationship between mental contamination and the core dimensions of contact contamination fear. J Anxiety Disord 45:9–16.

18. Olatunji BO, Liu Q, Knowles KA, Jessup SC (2025): State and Trait Disgust Uniquely Predict Avoidance in Contamination Fear: Specificity of Disease-Specific and Nonspecific Individual Differences. Behav Ther 56:32–42.

19. Bosman RC, Borg C, de Jong PJ (2016): Optimising Extinction of Conditioned Disgust. PLoS One 11:e0148626.

20. Ludvik D, Boschen MJ, Neumann DL (2015): Effective behavioural strategies for reducing disgust in contamination-related OCD: A review. Clin Psychol Rev 42:116–129.

21. Olatunji BO, Smits JAJ, Connolly K, Willems J, Lohr JM (2007): Examination of the decline in fear and disgust during exposure to threat-relevant stimuli in blood-injection-injury phobia. J Anxiety Disord 21:445–455.

22. Wang J, Becker B, Wang Y, Ming X, Lei Y, Wikgren J (2024): Conceptual-level disgust conditioning in contamination-based obsessive-compulsive disorder. Psychophysiology 61:e14637.

23. Ball TM, Gunaydin LA (2022): Measuring maladaptive avoidance: from animal models to clinical anxiety. Neuropsychopharmacology 47:978–986.

24. Endrass T, Kloft L, Kaufmann C, Kathmann N (2011): Approach and avoidance learning in obsessive-compulsive disorder. Depress Anxiety 28:166–172.

25. Provenza NR, Reddy S, Allam AK, Rajesh SV, Diab N, Reyes G, et al. (2024): Disruption of neural periodicity predicts clinical response after deep brain stimulation for obsessive-compulsive disorder. Nat Med 30:3004–3014.

26. Yu J, Zhou P, Yuan S, Wu Y, Wang C, Zhang N, et al. (2022): Symptom provocation in obsessive-compulsive disorder: A voxel-based meta-analysis and meta-analytic connectivity modeling. J Psychiatr Res 146:125–134.

27. Bhikram T, Abi-Jaoude E, Sandor P (2017): OCD: obsessive-compulsive … disgust? The role of disgust in obsessive-compulsive disorder. J Psychiatry Neurosci 42:300–306.

28. Liu J, Wang J, Song Y, Becker B, Ming X, Lei Y (2025): Enhanced disgust generalization in obsessive-compulsive disorder is related to insula and putamen hyperactivity. Psychol Med 55:e116.

29. Salmani B, Mancini F, Hasani J, Zanjani Z (2022): Anti-disgust cognitive behavioral therapy for contamination-based obsessive compulsive disorder: a randomized controlled clinical trial. J Clin Med 11:2875.

30. Mantione M, Nieman DH, Figee M, Denys D (2014): Cognitive-behavioural therapy augments the effects of deep brain stimulation in obsessive-compulsive disorder. Psychol Med 44:3515–3522.

31. Rodriguez-Romaguera J, Do Monte FHM, Quirk GJ (2012): Deep brain stimulation of the ventral striatum enhances extinction of conditioned fear. Proc Natl Acad Sci U S A 109:8764–8769.

32. Hamilton M (1960): A rating scale for depression. J Neurol Neurosurg Psychiatry 23:56–62.

33. Storch EA, Rasmussen SA, Price LH, Larson MJ, Murphy TK, Goodman WK (2010): Development and psychometric evaluation of the Yale-Brown Obsessive-Compulsive Scale–Second Edition. Psychol Assess 22:223–232.

34. Shofty B, Gadot R, Viswanathan A, Provenza NR, Storch EA, McKay SA, et al. (2023): Intraoperative valence testing to adjudicate between ventral capsule/ventral striatum and bed nucleus of the stria terminalis target selection in deep brain stimulation for obsessive-compulsive disorder. J Neurosurg 139:442–450.

35. Cheer JF, Aragona BJ, Heien MLAV, Seipel AT, Carelli RM, Wightman RM (2007): Coordinated Accumbal Dopamine Release and Neural Activity Drive Goal-Directed Behavior. Neuron 54:237–244.

36. Haber SN, Knutson B (2010): The Reward Circuit: Linking Primate Anatomy and Human Imaging. Neuropsychopharmacology 35:4–26.

37. Botvinick MM, Cohen JD, Carter CS (2004): Conflict monitoring and anterior cingulate cortex: an update. Trends Cogn Sci 8:539–546.

38. Roelofs K (2017): Freeze for action: neurobiological mechanisms in animal and human freezing. Philos Trans R Soc Lond B Biol Sci 372:20160206.

39. Fajnerova I, Gregus D, Francova A, Noskova E, Koprivova J, Stopkova P, et al. (2020): Functional Connectivity Changes in Obsessive-Compulsive Disorder Correspond to Interference Control and Obsessions Severity. Front Neurol 11:568.

40. Gürsel DA, Avram M, Sorg C, Brandl F, Koch K (2018): Frontoparietal areas link impairments of large-scale intrinsic brain networks with aberrant fronto-striatal interactions in OCD: a meta-analysis of resting-state functional connectivity. Neurosci Biobehav Rev 87:151–160.

41. Milad MR, Rauch SL (2012): Obsessive-compulsive disorder: beyond segregated cortico-striatal pathways. Trends Cogn Sci 16:43–51.

42. Tyagi H, Apergis-Schoute AM, Akram H, Foltynie T, Limousin P, Drummond LM, et al. (2019): A Randomized Trial Directly Comparing Ventral Capsule and Anteromedial Subthalamic Nucleus Stimulation in Obsessive-Compulsive Disorder: Clinical and Imaging Evidence for Dissociable Effects. Biol Psychiatry 85:726–734.

43. Banca P, Voon V, Vestergaard MD, Philipiak G, Almeida I, Pocinho F, et al. (2015): Imbalance in habitual versus goal directed neural systems during symptom provocation in obsessive-compulsive disorder. Brain 138:798–811.

44. Kim T, Lee SW, Lho SK, Moon SY, Kim M, Kwon JS (2024): Neurocomputational model of compulsivity: deviating from an uncertain goal-directed system. Brain 147:2230–2244.

45. Sheth SA, Mayberg HS (2023): Deep Brain Stimulation for Obsessive-Compulsive Disorder and Depression. Annu Rev Neurosci 46:341–358.

46. Slepneva N, Basich-Pease G, Reid L, Frank AC, Norbu T, Krystal AD, et al. (2024): Therapeutic DBS for OCD Suppresses the Default Mode Network. bioRxiv 2024.07.21.601827.

47. Widge AS, Zorowitz S, Basu I, Paulk AC, Cash SS, Eskandar EN, et al. (2019): Deep brain stimulation of the internal capsule enhances human cognitive control and prefrontal cortex function. Nat Commun 10:1536.

48. Denys D, Graat I, Mocking R, de Koning P, Vulink N, Figee M, et al. (2020): Efficacy of Deep Brain Stimulation of the Ventral Anterior Limb of the Internal Capsule for Refractory Obsessive-Compulsive Disorder: A Clinical Cohort of 70 Patients. Am J Psychiatry 177:265–271.

49. Provenza NR, Sheth SA, Dastin-van Rijn EM, Mathura RK, Ding Y, Vogt GS, et al. (2021): Long-term ecological assessment of intracranial electrophysiology synchronized to behavioral markers in obsessive-compulsive disorder. Nat Med 27:2154–2164.

50. Holtzheimer PE, Kelley ME, Gross RE, Filkowski MM, Garlow SJ, Barrocas A, et al. (2012): Subcallosal cingulate deep brain stimulation for treatment-resistant unipolar and bipolar depression. Arch Gen Psychiatry 69:150–158.

51. Malone DA, Dougherty DD, Rezai AR, Carpenter LL, Friehs GM, Eskandar EN, et al. (2009): Deep Brain Stimulation of the Ventral Capsule/Ventral Striatum for Treatment-Resistant Depression. Biol Psychiatry 65:267–275.

52. Gadot R, Li N, Shofty B, Avendano-Ortega M, McKay S, Bijanki KR, et al. (2024): Tractography-Based Modeling Explains Treatment Outcomes in Patients Undergoing Deep Brain Stimulation for Obsessive-Compulsive Disorder. Biol Psychiatry 96:95–100.

53. Li N, Baldermann JC, Kibleur A, Treu S, Akram H, Elias GJB, et al. (2020): A unified connectomic target for deep brain stimulation in obsessive-compulsive disorder. Nat Commun 11:3364.

54. McGovern RA, Sheth SA (2017): Role of the dorsal anterior cingulate cortex in obsessive-compulsive disorder: converging evidence from cognitive neuroscience and psychiatric neurosurgery. J Neurosurg 126:132–147.

55. Duan Y, Ma Z, Tsai PJ, Lu H, Xiao X, Wang D, et al. (2025): Frontostriatal regulation of brain circuits contributes to flexible decision making. Neuropsychopharmacology 50:1156–1166.

56. Ghane M, Trambaiolli L, Bertocci MA, Martinez-Rivera FJ, Chase HW, Brady T, et al. (2024): Specific Patterns of Endogenous Functional Connectivity Are Associated With Harm Avoidance in Obsessive-Compulsive Disorder. Biol Psychiatry 96:137–146.

57. Angelakis I, Pseftogianni F (2021): Association between obsessive-compulsive and related disorders and experiential avoidance: A systematic review and meta-analysis. J Psychiatr Res 138:228–239.

58. McGuire JF, Storch EA, Lewin AB, Price LH, Rasmussen SA, Goodman WK (2012): The role of avoidance in the phenomenology of obsessive-compulsive disorder. Compr Psychiatry 53:187–194.

59. Wheaton MG, Gershkovich M, Gallagher T, Foa EB, Simpson HB (2018): Behavioral avoidance predicts treatment outcome with exposure and response prevention for obsessive-compulsive disorder. Depress Anxiety 35:256–263.

60. Eng GK, Collins KA, Brown C, Ludlow M, Tobe RH, Iosifescu DV, Stern ER (2022): Relationships between interoceptive sensibility and resting-state functional connectivity of the insula in obsessive-compulsive disorder. Cereb Cortex 32:5285–5300.

61. Viol K, Aas B, Kastinger A, Kronbichler M, Schöller HJ, Reiter EM, et al. (2019): Erroneously Disgusted: fMRI Study Supports Disgust-Related Neural Reuse in Obsessive-Compulsive Disorder (OCD). Front Behav Neurosci 13:81.

62. Fink J, Pflugradt E, Stierle C, Exner C (2018): Changing disgust through imagery rescripting and cognitive reappraisal in contamination-based obsessive-compulsive disorder. J Anxiety Disord 54:36–48.

63. Olatunji BO, Berg H, Cox RC, Billingsley A (2017): The effects of cognitive reappraisal on conditioned disgust in contamination-based OCD: An analogue study. J Anxiety Disord 51:86–93.

